# Brain iron and mental health symptoms in youth with and without prenatal alcohol exposure

**DOI:** 10.1101/2022.03.22.22272038

**Authors:** Daphne Nakhid, A. McMorris Carly, Hongfu Sun, William B. Gibbard, Christina Tortorelli, Catherine Lebel

**Affiliations:** Department of neuroscience, University of Calgary, Calgary, AB, T2N 1N4, Canada; Alberta Children’s Hospital Research Institute (ACHRI), University of Calgary, Calgary, AB, T2N 1N4, Canada; Hotchkiss Brain Institute, University of Calgary, Calgary, AB, T2N 1N4, Canada; Werklund School of Education, School and Applied Child Psychology; Alberta Children’s Hospital Research Institute (ACHRI); Department of Pediatrics, University of Calgary, Calgary, AB, T2N 1N4, Canada; School of Information Technology and Electrical Engineering, University of Queensland, Brisbane, 4072, Australia; Department of Child Studies and Social Work, Mount Royal University, Calgary, Alberta, T3E 6K6, Canada; Department of Radiology; University of Calgary, Calgary, AB, T2N 1N4, Canada

**Keywords:** prenatal alcohol exposure, FASD, brain, iron, QSM, MRI, mental health, internalizing problems, externalizing problems, youth

## Abstract

Prenatal alcohol exposure (PAE) negatively affects brain development and increases the risk of poor mental health. We investigated if brain susceptibility, measuring iron, or volume were associated with internalizing or externalizing symptoms in youth with and without PAE. T1-weighted and quantitative susceptibility mapping (QSM) MRI scans were collected for 19 PAE and 40 unexposed participants aged 7.5-15 years. Magnetic susceptibility and volume of basal ganglia and limbic structures were extracted using FreeSurfer. Internalizing and externalizing problems were assessed using the Behavioural Assessment System for Children (BASC-2-PRS). Susceptibility in the nucleus accumbens was negatively associated with internalizing problems, while amygdala susceptibility was positively associated with internalizing problems across groups. In the PAE group, thalamus susceptibility was negatively associated with internalizing problems, and putamen susceptibility was positively associated with externalizing problems. Brain volume was not related to internalizing or externalizing symptoms. These findings highlight that brain iron is related to internalizing and externalizing symptoms differently in some brain regions for youth with and without PAE. Atypical iron levels (high or low) may indicate mental health problems across individuals, and iron in the thalamus and putamen may be particularly important for behaviour in individuals with PAE.

## Introduction

Prenatal alcohol exposure (PAE) can affect fetal development, resulting in altered neuroanatomy, neurophysiology, and increasing the risk of poor mental health outcomes. Fetal alcohol spectrum disorder (FASD) is the neurodevelopmental disability associated with PAE, and the most common preventable cause of developmental disabilities in children [1–3]. PAE is linked to aberrant structural and functional brain development, including globally reduced brain volume, reduced white and grey matter volume (see reviews [4-5]), altered cortical thickness [6], altered white matter connectivity [7-8], and abnormal brain function [9].

PAE increases the risk of poor mental health outcomes; approximately 90% of individuals with FASD have mental health problems [10]. Evidence of mental health problems in infants with PAE can be observed as early as 2 years of age [11] and continues to increase through childhood and adolescence, persisting into adulthood [10]. Children with PAE experience more mental health problems than their ability-matched peers [12]. Mental health problems in children can be thought of as internalizing (negative behaviours directed inwards), such as anxiety or depression, and externalizing (negative behaviours directed outwards), such as aggression and hyperactivity. Internalizing disorders such as anxiety and depression frequently co-occur with FASD and are 4 and 11 times more common than in the general population [13]. Similarly, externalizing disorders such as attention-deficit/hyperactivity disorder (ADHD) and oppositional defiant disorder are 10 and 13 time more common than in the general population [13]. Unexposed youth and adults with internalizing (depression and anxiety) and externalizing (ADHD) disorders tend to show volumetric abnormalities of limbic and basal ganglia structures [14–22]. Youth with PAE have similar volumetric abnormalities [23–25]; however, little research has investigated the neural underpinnings between brain and internalizing or externalizing symptoms in individuals with PAE.

In the general population, brain iron, and in particular, iron deficiency anemia, has been linked to developmental disabilities and mental health problems in youth [26] and adults [27]. Early brain development requires iron for myelination [28-29], monoamine synthesis, and neurotransmitter metabolism [29-30]. In the first six months of life, infants depend on fetal iron stores accumulated during the third trimester of pregnancy [31-32]. Infants and children who have an iron deficiency show increased cognitive and affective problems (e.g., less social smiling and looking) [33-34]. Altered developmental and mental health outcomes associated with iron deficiency persist for years even after treatment [35], highlighting the importance of sufficient iron availability throughout development.

Literature investigating brain iron in individuals with PAE is very limited. Prenatal maternal binge drinking increases the risk of iron deficiency anemia in infants at 12 months of age [31]. In animal models, PAE lowers fetal brain iron [36] and behavioural deficits persist even after iron levels normalize [37]. Blood iron deficiency in infancy is related to emotional withdrawal, which predicts affective problems in childhood [38]. Of note, the relation between blood iron and brain iron is not well understood, and blood iron does not always correlate with brain iron, especially in children with neurodevelopmental disabilities [39]. In other neurodevelopmental disabilities, preschoolers with autism have lower brain iron in the caudate nucleus [40] and children with ADHD have lower brain iron in the caudate, putamen, and thalamus compared to controls [39-41]. Although brain iron is associated with poor developmental and mental health outcomes, issues that are experienced by most individuals with PAE, no studies to date have looked at brain iron in youth with PAE, and how it is associated with internalizing and externalizing issues.

Transverse relaxation rates (*R*2*), and susceptibility weighted imaging (SWI) have both been validated as correlated to iron concentration [42–45], though they are indirectly related to brain iron and lack specificity [46-47]. Quantitative susceptibility mapping (QSM) measures tissue susceptibility, an intrinsic property that determines how tissue will interact with a magnetic field and provides a more reliable, reproducible, and valid measure of brain iron. Iron is paramagnetic (positive susceptibility) and has been validated post-mortem as the primary susceptibility source in subcortical gray matter [48-49].

Here, we examined the association between brain iron and internalizing or externalizing symptoms in youth and determined how group members (exposed versus unexposed) moderates or impacts this association. Consistent with previous literature, we focused on key brain regions: the basal ganglia (nucleus accumbens, caudate, putamen and pallidum), a primary site of iron accumulation, as well as limbic structures (thalamus, hippocampus, and amygdala) which have been implicated in mental health issues. Based on previous research on brain iron in children with externalizing symptoms [39-41] and adults with internalizing symptoms [50-51], it was expected that across diagnostic groups, lower susceptibility of the caudate and putamen would be associated with more internalizing and externalizing symptoms and a group-susceptibility interaction was anticipated, such that in the PAE group lower susceptibility in the thalamus would be associated with more internalizing and externalizing symptoms. Consistent with Krueger and colleagues who found a trend-level association with caudate volume and internalizing symptoms in children with PAE [52], we hypothesized that the PAE group would have lower caudate volume and that this would be associated with more internalizing symptoms. It was anticipated that participant gender and age would also influence these associations.

## Methods

### Participants

#### PAE group

Nineteen youth with PAE (11.18 ± 2.16; 10M/9F) aged 7.5-15 years were recruited through posters, social media, word of mouth, caregiver support groups, early intervention services, and the Cumulative Risk Diagnostic Clinic in Calgary, Alberta (Canada). PAE was confirmed through child welfare files, medical, police, and social work records (when available), as well as semi-structured interviews with caregivers and/or birth families. Thirteen participants with PAE had confirmed alcohol exposure meeting the Canadian diagnostic guidelines for FASD (≥7 drinks/week or ≥2 binge episodes of at least 4 drinks at some point in pregnancy) and six participants with PAE had confirmed exposure of an unknown amount. Seven participants had a diagnosis of FASD, and one participant was classified as at risk of developing FASD. Some participants with PAE had a prior diagnosis of ADHD (n=8), learning disability (n=1), anxiety (n=1), attachment disorder (n=2), borderline intellectual functioning (n=1), mild mental disability (n=1), oppositional defiant disorder (n=1); three participants with PAE had more than one diagnosis. Ten participants were taking medication (seven for ADHD symptoms, three for other symptoms) at the time of their scan. No participants had MRI contraindications. Gender was parent/caregiver reported.

#### Unexposed group

Forty unexposed youth (11.18 ± 2.27; 23M/17F) 7.5-15 years of age were recruited through posters, social media, and word of mouth. These participants had confirmed absence of PAE based on retrospective questionnaires completed by a caregiver at the time of the MRI scan. These participants had no prior or suspected diagnosis of a neurodevelopmental disability or mental health issues, and none of these participants were taking medication related to developmental and mental health difficulties. Participants had no MRI contraindications and gender was parent/caregiver reported.

### MRI acquisition

Participants completed an MRI scan on a 3T GE MR750w MRI scanner at the Alberta Children’s Hospital using a 32-channel head coil. The imaging protocol included a T1-weighted anatomical scan acquired using a 3D FSPGR sequence with TI = 600 ms, TR = 8.2 ms, 0.8 mm isotropic resolution and total scan time of 5:38 minutes. QSM images were acquired using a 3-dimensional spoiled gradient recalled sequence with the acquisition parameters: repetition time (TR) = 56.8 ms, Echo Time (TE) = 4.5 ms to 52.2 ms for a total of 10 echoes with 5.3 ms spacing between each echo, field of view = 240×240×182.4 mm^3^, acquired voxel size = 0.94×1.17×1.90 mm^3^, reconstructed voxel size = 0.94×0.94×1.90 mm^3^, flip angle = 10°, acceleration factor = 2.5, total scan time of 5:14 minutes.

### T1-weighted image processing

FreeSurfer 6.0’s automated recon-all pipeline was used for processing, editing, and segmenting the T1-weighted anatomical images [53-54]. The recon-all pipeline registers participants’ brains to a template brain, performs skull stripping, image registration, intensity normalization, segmentation, and performs volume calculations. Segmentations were manually checked to ensure proper segmentation of the outer pial and white matter border, and manual editing was performed as necessary. The segmentations from FreeSurfer were used to extract the bilateral caudate, putamen, pallidum, thalamus, amygdala, hippocampus, and nucleus accumbens (Figure 1).

**Figure 1:**
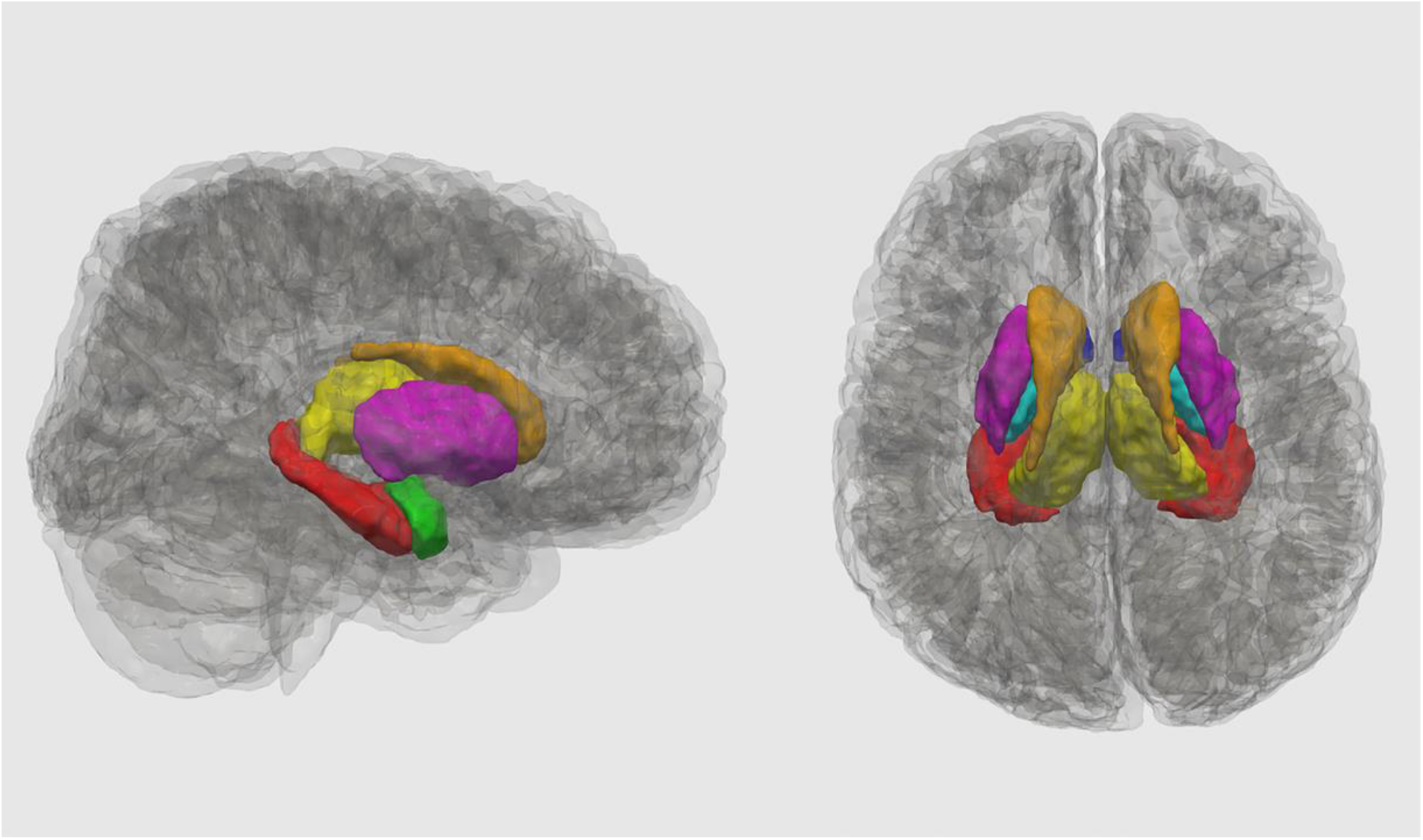
Regions of interest in an unexposed control participant (girl, age range 7-15) in a sagittal (left) and axial (right) 3D mesh reconstruction using ITK-Snap: nucleus accumbens (dark blue), caudate (copper), putamen (pink), pallidum (cyan), thalamus (yellow), hippocampus (red) and amygdala (lime green).

### QSM reconstruction

The first five echoes (TE from 4.5 ms to 25.7 ms) were used for QSM reconstruction. The raw phase was unwrapped using a best-path method [55], RESHARP [56] was used for background field removal, and iLSQR [57] was used for dipole inversion. The QSM processing pipeline was implemented in MATLAB [58] source code is available on GitHub: https://github.com/sunhongfu/QSM. Mean susceptibility of the whole brain was used as a point of reference since there is no susceptibility bias during analysis whether referencing the whole brain or a specific region [59–61]. Segmentations from FreeSurfer were transformed to QSM space using mri_robust_register [62] and were used to extract the susceptibility values of the caudate, putamen, pallidum, thalamus, amygdala, hippocampus, and nucleus accumbens bilaterally.

### Behavioural Measures

Caregivers completed the Behavioural Assessment System for Children, Second Edition–Parent Rating Scale (BASC-2-PRS; [63]) to measure internalizing (e.g., anxiety, depression, somatization) and externalizing symptoms (aggression, conduct, and hyperactivity). The BASC-2 provides *T* scores that are age and gender normed; higher scores indicate worse symptoms. Scores <60 are considered developmentally appropriate, scores of 60-69 are considered at-risk of developing a disorder, and scores ≥70 are considered clinically maladaptive behaviour indicating that a child may meet diagnostic criteria for a given internalizing or externalizing disorder. The Internalizing Problems composite score includes anxiety, depression, and somatization subscales while the Externalizing Problems composite score includes aggression, conduct problems, and hyperactivity subscales. The *T* score for the Internalizing Problems and Externalizing Problems was used in all analyses, unless otherwise specified.

### Statistical analysis

R Statistics version 4.0.0 [64] was used for all analyses. To determine if groups differed by age and gender, a chi-square analysis (group by gender) and independent sample *t-*test (group by age) was conducted. To determine if diagnostic groups differed on internalizing and externalizing symptoms, a two-sample Wilcoxon rank test was used. To determine if there was a main effect of group (PAE and unexposed), brain measures (susceptibility/ brain iron or volume) or a group-brain interaction, two-way ANCOVAs for each brain region were conducted to determine the association between internalizing or externalizing problems and volume or susceptibility by group, with age and gender as covariates. If the group-brain interaction was not significant, it was removed from the model. For structures where susceptibility or volume was significantly associated with behaviour, we also tested associations using a two-way ANCOVA with internalizing (depression, anxiety, somatization) or externalizing (hyperactivity, conduct problems, aggression) subscales. Results are reported both uncorrected (*p*<.05) and corrected for multiple comparisons using Benjamini-Hochberg false discovery rate (FDR) at *q*<.05.

## Results

### Demographics

There were no group differences in gender (*X*^2^(1, N=59)=.124, *p*=.725) or age (*t* (37.057)=-0.005, *p*=.996).

### Behavioural outcomes

The median externalizing *T* score in the PAE group was 68 (IQR=10), whereas the median in unexposed group was 50 (IQR=9.25), this difference was significant (*p*<0.001; Table 1). The median aggression *T* score in PAE group was 60 (IQR = 11), whereas the median in unexposed group was 50 (IQR=8.25), this difference was significant (*p*<0.001). The median conduct *T* score in PAE group was 60 (IQR=15.5), whereas the median in unexposed group was 51 (IQR=14), this difference was significant (*p*=0.002). The median hyperactivity *T* score in PAE group was 69 (IQR=13.5), whereas the median in unexposed group was 50 (IQR = 10), this difference was significant (*p*<0.001). There were no significant differences in internalizing symptoms by diagnostic group.

**Table 1.** Comparison of internalizing Behavioural Assessment System for Children (BASC-2-PRS) *T* scores by diagnostic group.

|  |  | **Diagnostic group** |  | **Wilcoxon rank test** |
| --- | --- | --- | --- | --- |
|  |  | Prenatal alcohol exposure (PAE) (n=19) | Unexposed Controls (n=40) |  |
| | Mean Score ± SD | 57 ± 19 | 51 ± 11 | $W=423, p = .490$ |
| **Internalizing problems composite score** | At risk (60-69) | 2 (11%) | 3 (7.5%) |  |
| | Clinically significant $\geq 70$ | 4 (21%) | 4 (10%) | |
|  | Total | 6 (32%) | 7 (17.5%) |  |
| **Anxiety subscale** | Mean Score ± SD | 51 ± 14 | 51 ± 13 | W=372, $p=.900$ |
|  | At risk (60-69) | 5 (26%) | 3 (8%) |  |
| | Clinically significant $\geq 70$ | 2 (11%) | 5 (13%) | |
|  | Total | 7 (37%) | 8 (20%) |  |
| **Depression subscale** | Mean Score ± SD | 58 ± 15 | 52 ± 11 | W=477, $p=.119$ |
|  | At risk (60-69) | 4 (21%) | 3 (8%) |  |
| | Clinically significant $\geq 70$ | 4 (21%) | 4 (10%) | |
|  | Total | 8 (42%) | 7 (18%) |  |
| **Somatization subscale** | Mean Score ± SD | 58 ± 22 | 49 ± 10 | W=428, $p=.445$ |
|  | At risk (60-69) | 1 (5%) | 6 (15%) |  |
| | Clinically significant $\geq 70$ | 5 (26%) | 1 (3%) | |
|  | Total | 6 (32%) | 7 (18%) |  |

\* $p < .05$
\*\* $p \leq .001$

**Table 2.** Comparison of externalizing BASC-2-PRS *T* scores by diagnostic group

|  |  | **Diagnostic group** |  | **Wilcoxon rank test** |
| --- | --- | --- | --- | --- |
|  |  | PAE (n=19) | Unexposed Controls (n=40) |  |
| **Externalizing problems composite score** | Mean Score ± SD | 63 ± 12 | 53 ± 9 | W=604, $p < .001^{**}$ |
|  | At risk (60-69) | 9 (47%) | 4 (10%) |  |
| | Clinically significant $\geq 70$ | 5 (26%) | 2 (5%) | |
|  | Total | 14 (73%) | 6 (15%) |  |
| **Aggression subscale** | Mean Score ± SD | 59 ± 8 | 51 ± 8 | W=604, $p < .001^{**}$ |
|  | At risk (60-69) | 8 (42%) | 2 (5%) |  |
| | Clinically significant $\geq 70$ | 2 (11%) | 2 (5%) | |
|  | Total | 10<br>(53%) | 4 (10%) |  |
| **Conduct subscale** | Mean Score ± SD | 63 ± 13 | 52 ± 10 | $W=575$,<br>$p=.002^*$ |
|  | At risk (60-69) | 5 (26%) | 5 (12.5%) |  |
| | Clinically significant $\geq 70$ | 5 (26%) | 3 (7.5%) | |
|  | Total | 10<br>(52%) | 8 (20%) |  |
| **Hyperactivity subscale** | Mean Score ± SD | 66 ± 11 | 52 ± 11 | $W=626$,<br>$p<.001^{**}$ |
|  | At risk (60-69) | 8 (42%) | 5 (12.5%) |  |
| | Clinically significant $\geq 70$ | 6 (32%) | 2 (5%) | |
|  | Total | 14<br>(74%) | 8 (17.5%) |  |

\* $p<.05$
\*\* $p\leq.001$

### Magnetic susceptibility and behavioural outcomes

Across groups, susceptibility of the nucleus accumbens was negatively associated with internalizing *T* scores (*F*(1, 54)=7.020, *p*=.011, *q*=.056), as well as depression (*F*(1, 54)=5.447, *p*=.023, *q*=.069) and anxiety (*F*(1, 54)=12.455, *p*<.001, *q*=.003) subscales (Figure 2). Amygdala susceptibility was positively associated with internalizing *T* scores (*F*(1, 54)=4.168, *p*=.046, *q*=.107) and the anxiety subscale (*F*(1, 54)=5.009, *p*=.029, *q*=.029; Figure 3).

**Figure 2.**
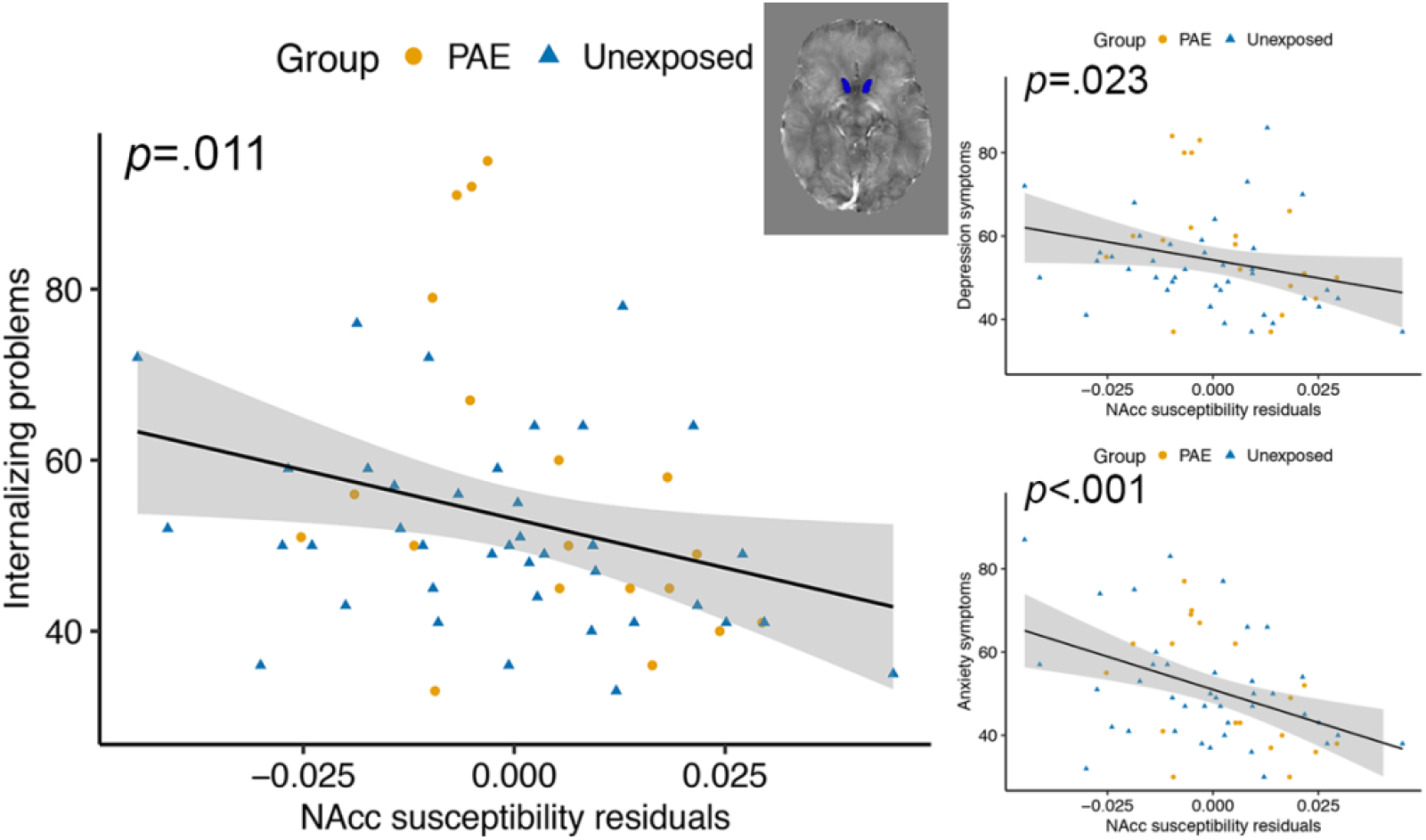
Nucleus accumbens (NAcc) magnetic susceptibility was negatively associated with BASC-2-PRS *T* scores internalizing problems, depression, and anxiety symptoms in youth with and without PAE. The *p* values are uncorrected, and susceptibility values are corrected for age and gender.

**Figure 3.**
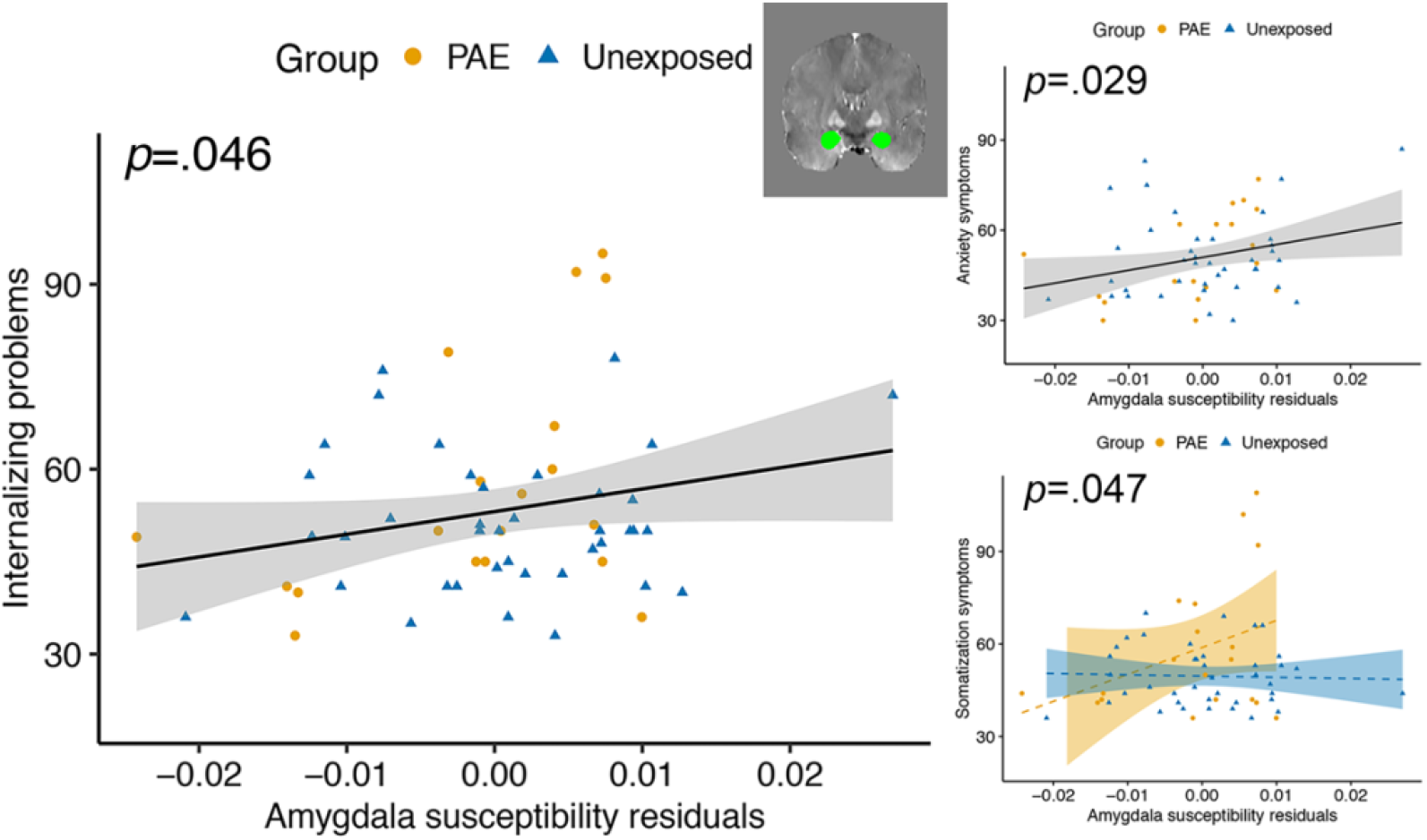
Amygdala magnetic susceptibility was positively associated with BASC-2-PRS *T* scores for internalizing problems and anxiety symptoms in youth with and without PAE. Group-susceptibility interactions in the amygdala showed higher susceptibility was associated with more somatization symptoms in youth with PAE and higher susceptibility was associated with fewer somatization symptoms in unexposed youth. The *p* values are uncorrected, and susceptibility values are corrected for age and gender. Dashed lines indicate non-significant associations within a group for amygdala susceptibility.

There was a group-susceptibility interaction for the thalamus for internalizing *T* scores (*F*(1, 53)=6.157, *p*=.016, *q*=.056), anxiety (*F*(1, 53)=9.305, *p*=.004, *q*=.006), and depression (*F*(1, 53)=4.043, *p*=.049, *q*=.074), such that scores were negatively correlated with susceptibility for the PAE group and positively correlated in controls (Figure 4). There was a group-susceptibility interaction for the somatization subscale (*F*(1, 53)= 4.121, *p*=.047, *q*=.141), such that there was a positive association between amygdala susceptibility and somatization symptoms in the PAE group and a slightly negative association in unexposed participants (Figure 3).

**Figure 4.**
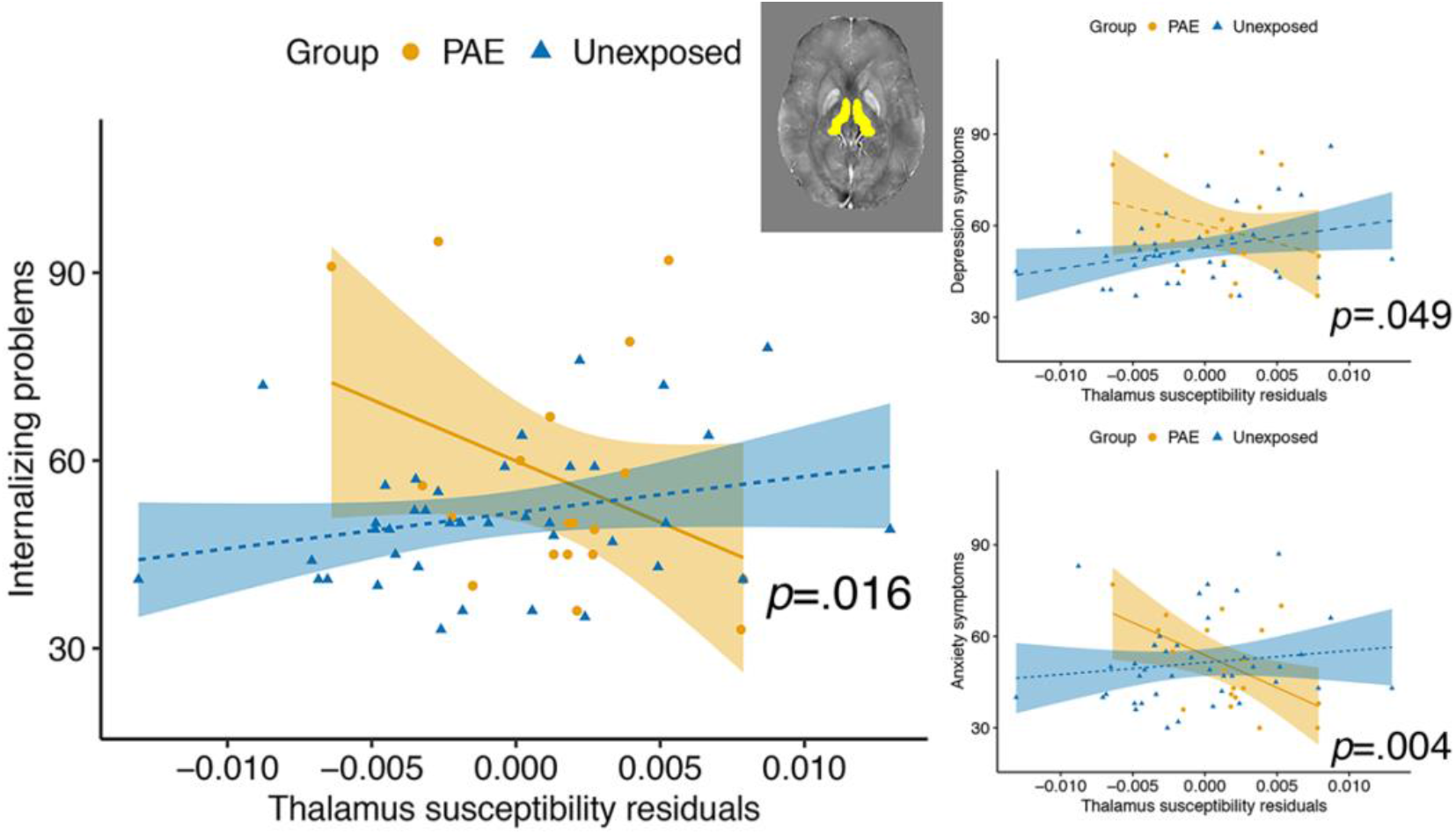
Group-susceptibility interactions in the thalamus showed that higher susceptibility is associated with BASC-2-PRS *T* scores indicating fewer internalizing problems, depression, and anxiety symptoms in youth with PAE. Higher thalamus magnetic susceptibility was associated with more internalizing problems, depression, and anxiety symptoms in unexposed youth. The *p* values are uncorrected, and susceptibility values are corrected for age and gender. Dashed lines indicate non-significant associations within a group for thalamus susceptibility.

There was a group-susceptibility interaction for the putamen for externalizing problems (*F*(1, 53)=4.497, *p*=.039, *q*=.273) with the PAE group showing more externalizing problems associated with higher susceptibility and the unexposed group having a slightly negative correlation (Figure 5). None of the externalizing subscales was significantly related to putamen susceptibility.

**Figure 5.**
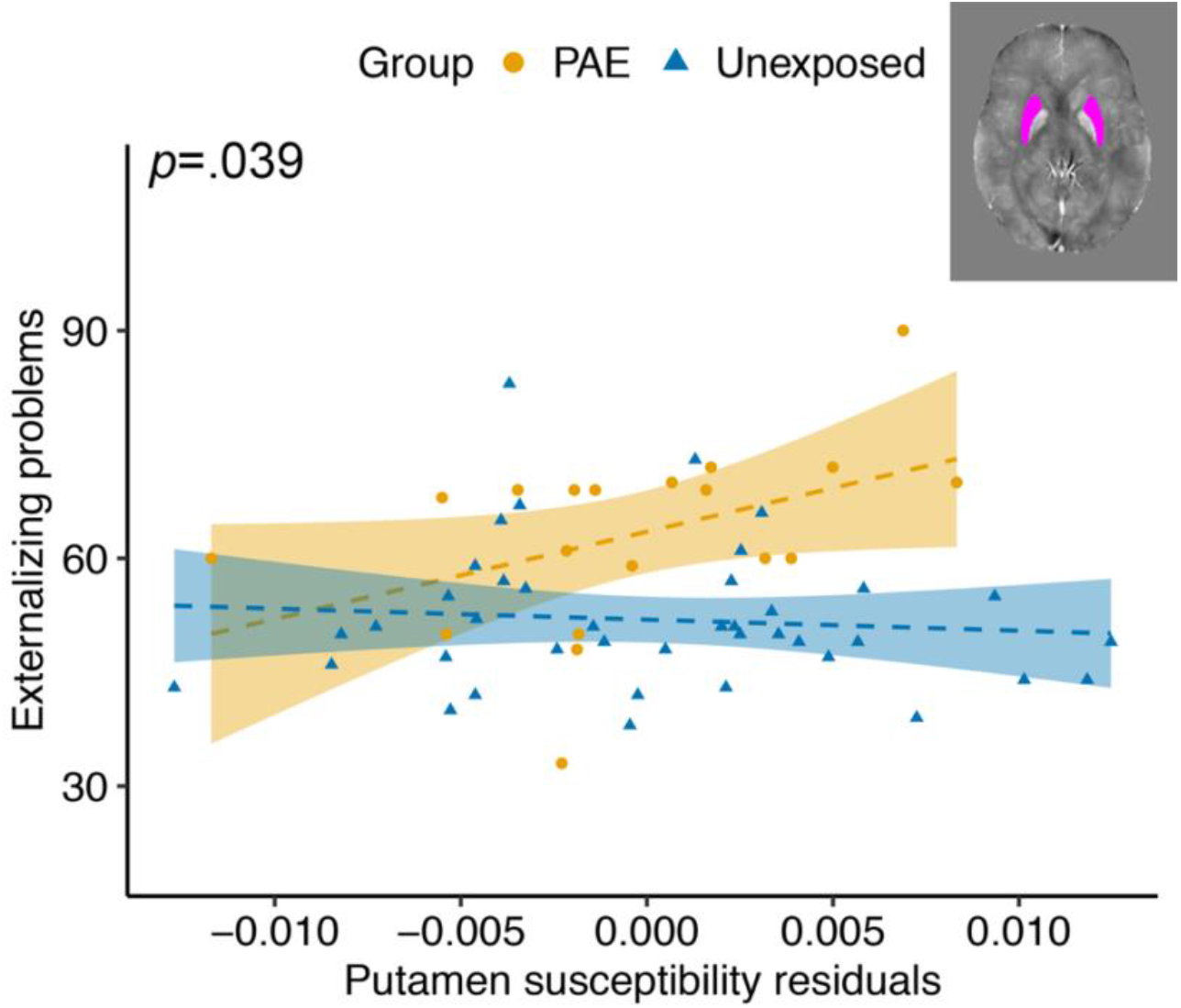
Group-susceptibility interactions in the putamen show that higher susceptibility is associated with BASC-2-PRS *T* scores for externalizing problems in youth with PAE. Higher susceptibility is associated with less externalizing problems in unexposed youth. The *p* values are uncorrected, and susceptibility values are corrected for age and gender. Dashed lines indicate non-significant associations within a group for putamen susceptibility.

### Brain volume and behavioural outcomes

There were no brain volume or group-volume interactions associated with internalizing or externalizing problems.

## Discussion

For the first time, we showed that brain iron is associated with internalizing and externalizing problems in youth, and in some cases was moderated by PAE. Across groups, less brain iron (lower susceptibility) in the nucleus accumbens was associated with more internalizing problems, while in the amygdala, higher brain iron (greater susceptibility) was associated with more internalizing problems. This suggests that the effects of brain iron on mental health symptoms differ regionally. Our findings in the thalamus and putamen demonstrate that PAE not only alters brain iron, but also disrupts the brain-behaviour relationship. No associations between brain volumes and mental health symptoms were found, suggesting that more specific measures such as susceptibility may be helpful in determining what may be contributing to mental health problems in this population.

As expected, the PAE group had significantly more externalizing, but not internalizing, symptoms than unexposed controls. Externalizing symptoms are well-recognized in individuals with PAE; ADHD (an externalizing disorder) occurs 10 times more often in people with PAE than those unexposed and it is the most common co-occurring disorder with FASD [13]. Many studies report that externalizing symptoms are more affected by PAE than internalizing symptoms at this age [12]. However, internalizing symptoms can be hard to measure with parent report since internalizing symptoms are not as easily observed, which may be contributing to an under-reporting of internalizing symptoms across one or both groups.

Iron is an essential nutrient for proper brain development and behaviour. Alcohol consumption while pregnant can affect fetal iron stores [36], disrupting postnatal brain development. The nucleus accumbens plays an important role in motivation and emotional processing [65], and is implicated in a number of psychological disorders including depression and anxiety [66]. Based on previous research, we had no hypothesis about brain iron in the nucleus accumbens and internalizing or externalizing symptoms but found it was associated with internalizing problems, driven mostly by anxiety symptoms. The nucleus accumbens is part of the mesolimbic dopaminergic pathway, so low iron may impact behaviour through dopamine metabolism. Iron deficient rats show decreased dopamine transporter (DAT) in the striatum and nucleus accumbens, which can result in elevated dopamine leading to desensitization of the DAT [67]. Similarly, Youdim [68] found decreased dopamine 2 (D2) receptors in the nucleus accumbens of iron deficient rats, D2 receptors reduce acetylcholine release [69], the main neurotransmitter of the parasympathetic nervous system.

The amygdala plays an important role in limbic circuits and emotional responses including fear and anxiety [70]. Amygdala iron was positively related to internalizing problems and anxiety symptoms in youth with and without PAE. We also found that higher amygdala iron was associated with more somatization symptoms in the PAE group but not the unexposed controls. Inhibitory circuits in the amygdala, that are highly dependent on GABA, are important for promoting and suppressing anxiety-like behaviour [71]. The amygdala tends to have stable, low levels of brain iron across the lifespan [72]. It is possible that small perturbations in brain iron have a large effect on GABA metabolism and therefore on anxiety-like behaviour in this brain region.

The thalamus is involved in limbic, cognitive, and sensorimotor circuits and acts primarily as a relay station [73]. Lower thalamus susceptibility was associated with internalizing symptoms (internalizing problems, depression, and anxiety subscales). The lack of relationships with externalizing symptoms is surprising given that prior studies report lower thalamus iron in youth with ADHD [39,74], an externalizing disorder that often co-occurs with PAE. That being said, participants in this study with ADHD were almost all taking psychostimulant medication, which is thought to normalize brain iron in the thalamus [75]. Previous research in adults show increased brain iron in the thalamus and putamen is related to depression severity [50-51]. We saw evidence of this trend in the unexposed group that had increasing internalizing symptoms with higher brain iron, while the opposite relationship was observed in youth with PAE. The thalamus is rich in dopamine [76] and behavioural outcomes related to brain iron in the thalamus are likely related to dopamine metabolism.

The putamen is part of the striatum involved in reward, cognition, language, and addiction [77]. We hypothesized that lower susceptibility in the putamen would be associated with more internalizing and externalizing problems but found the opposite to be true. Previous research suggests that brain iron in the putamen is lower in children with ADHD [39]; however, in our sample, more brain iron in the putamen was associated with more externalizing symptoms in the PAE group. Previous research have found that iron in the putamen is related to depression severity in adults [50], but we did not find this in our sample. The putamen undergoes prolonged development [78], and it is possible that the effects of susceptibility on the putamen and behaviour would be better captured in older adolescence or young adulthood. Although the mechanism of how brain iron is related to internalizing and externalizing symptoms is not well understood, it is likely due to dopamine, serotonin, and norepinephrine metabolism [79].

In contrast to the QSM results, there were no associations between brain volume and behaviour. This differs from research on mental health and brain structure in unexposed participants that has found associations between caudate volume, putamen development, amygdala, hippocampus, and thalamus abnormalities and internalizing and externalizing disorders [14–22]. Only one study on children with PAE found a trend-level association between caudate volume and internalizing symptoms [52]. Another study, with an overlapping sample with our study, found no associations between brain volumes and behaviour [80]. Similarly, we found no associations between brain volumes and mental health symptoms. One explanation is that volume reductions in children with PAE may disrupt the etiology of mental health problems that are seen in unexposed individuals. However, further research to clarify these relationships is needed.

Nutritional interventions, such as choline supplementation, show promise for assisting healthy neurodevelopment in infants and children with PAE [81–83]. This opens the question of whether iron supplementation may be a viable intervention for individuals with PAE. Animal research has been inconclusive whether iron supplementation can reverse the effects of iron deficiency during development [37,84], indicating that timing of iron supplementation may be a critical factor. Future studies with prospective and large longitudinal samples are required to fully understand the impact of PAE on brain iron to assess iron supplementation as a potential nutritional intervention.

### Limitations

This is the first study to examine brain iron and mental health in children and youth with PAE; however, some limitations in the study design and methodology exist. Given our small sample, this study had limited power. Information about PAE was collected as rigorously as possible, however, given the retrospective nature of the study, details around exposure amounts, trimester, and frequency were not always available. Youth with PAE are at a greater risk of experiencing other prenatal exposures (e.g., tobacco, cocaine, marijuana; [85-86]) and postnatal adversities (e.g., neglect, deprivation, physical abuse, and caregiver changes; [85,87]; however, were not considered in our analyses. Considering that prenatal exposures and postnatal adversities negatively impact brain development [80,85], these should be controlled in future studies. Lastly, mental health symptoms were measured using caregiver report which is less reliable when measuring internalizing symptoms [88] and may be underestimating internalizing symptoms. Given the cognitive and emotional challenges youth with PAE face, caregiver report was chosen to be the best method for assessment collection; however, future studies should use both parent and self-report outcome measures when possible.

## Conclusions

Expanding on animal research, we show that brain iron is associated with internalizing symptoms, mainly anxiety, and externalizing symptoms, in multiple brain regions in youth with and without PAE. Group-brain interactions in internalizing and externalizing symptoms in some brain regions suggest that the effect of brain iron impacting behaviour differs between those with and without PAE. Our lack of findings between brain volume and internalizing or externalizing symptoms suggest a need for more specific brain measures, such as QSM used here, to better understand the neurological underpinnings of how PAE affects the brain and behaviour.

## Data Availability

Unexposed participant T1-weighted neuroimaging data is available at: https://figshare.com/articles/dataset/Structural_and_diffusion-weighted_images_of_the_
adolescent_brain/6002273 . PAE data is available upon request to the principal investigator.

https://figshare.com/articles/dataset/Structural_and_diffusion-weighted_images_of_the_adolescent_brain/6002273

## Author contributions

DN drafted and revised the manuscript, conducted the neuroimaging and statistical analyses, analyzed, and interpreted the data. CM, WBG, and CT, contributed to study concept and design, and revising the manuscript. HS wrote the code used for QSM analyses and critically revised the manuscript. CL led study concept and design, data interpretation, and revised the manuscript.

## Institutional review board statement

This study was approved by the University of Calgary Conjoint Health Research Ethics Board (REB 17-0663 approved June 8, 2017).

## Informed consent statement

Written informed consent was obtained from the parents and assent was obtained from the participants from all participants in this study.

## Data availability statement

Unexposed participant T1-weighted neuroimaging data is available at: https://figshare.com/articles/dataset/Structural_and_diffusion-weighted_images_of_the_adolescent_brain/6002273. PAE data is available upon request to the principal investigator.

